## Supplementary Material for "HepatoPredict accurately selects hepatocellular carcinoma patients for liver transplantation regardless of tumor heterogeneity"

**Tables**

**Supplementary Table 1 – Description of the currently used clinical criteria to select patients HCC for liver transplantation that were assessed in the study.**

| **Criteria** | **Description** |
| --- | --- |
| **Milan** ^1^ | 1 tumor with diameter ≤ 5 cm or ≤ 3 tumors each with diameter ≤ 3 cm, and no macrovascular invasion. |
| **UCSF** ^2^ | 1 tumor with diameter ≤ 6.5 cm, or ≤ 3 nodules with the largest lesion with diameter ≤ 4.5 cm and total tumor diameter ≤ 8 cm. |
| **AFP Score** ^3^ | Largest tumor diameter: ≤ 3 cm [0 points]; 3 - 6 cm [1 point]; > 6 cm [4 points] + Number of tumors: 1 - 3 [0 points]; ≥ 4 [2 points] + AFP level: ≤ 100 ng/mL [0 points]; 100 – 1000 ng/mL [2 points]; > 1000 ng/mL [3 points]. Score ≤ 2. |
| **ArgScore** ^4^ | AFP > 100 ng/ml [Yes = 1 point, No = 0 point], tumor beyond Up-to-7 [Yes = 1 point, No = 0 point}. Score = 0 points [low risk]. |
| **Warsaw** ^5^ | Expansion of Milan criteria including cases outside Milan criteria but within UCSF or Up to Seven (Up7) criteria with AFP < 100 ng/mL. |
| **MT2.0** ^6^ | Number of tumors and largest tumor diameter ≤ 7 + AFP < 200 ng/mL; Number of tumors and largest tumor diameter ≤ 5 cm + AFP < 400 ng/mL; Number of tumors and largest tumor diameter ≤ 4 cm + AFP < 1000 ng/mL. |
| **wALL** ^7^ | Intersection of the AFP Score and MT2.0. |

**Supplementary Table 2 – Performance metrics of the retrained HepatoPredict algorithm comparing to its previous version**^8^**.** The number of patients used to train each algorithm is indicated (n).

|  | **Sensitivity**  **(recall)** | **PPV**  **(precision)** | **Specificity** | **NPV** | **Accuracy** |
| --- | --- | --- | --- | --- | --- |
| **HepatoPredict previous version (n=162)** | | | | | |
| **HP Class I** | 0.48 | 0.95 | 0.92 | 0.34 | 0.58 |
| **HP Class II** | 0.43 | 0.73 | 0.46 | 0.19 | 0.44 |
| **HP Class I+II** | 0.91 | 0.83 | 0.38 | 0.56 | 0.79 |
| **HepatoPredict current version (n=232)** | | | | | |
| **HP Class I** | 0.75 | 0.92 | 0.76 | 0.46 | 0.75 |
| **HP Class II** | 0.96 | 0.86 | 0.44 | 0.77 | 0.85 |

**Supplementary Table 3 – Performance metrics/accuracy measures of the retrained HepatoPredict algorithm and other currently used clinical criteria in the testing subsets.** Data represented as mean ± SD. The retrained HepatoPredict (HP) was compared with Milan criteria (Milan) and the University of San Francisco California (UCSF) criteria (n = 69), whereas the HP AFP samples subset was compared with AFP-based criteria such as AFP score, metroticket 2.0 (MT2.0), Argentinian Score (ArgScore), Warsaw criteria (Warsaw), and within all criteria (wALL) (n = 42). rHP Class I is a subset of rHP Class II.

|  | **Sensitivity**  **(recall)** | **PPV**  **(precision)** | **Specificity** | **NPV** | **Accuracy** | **TP** | **FP** | **TN** | **FN** |
| --- | --- | --- | --- | --- | --- | --- | --- | --- | --- |
| **ALL SAMPLES** |  |  |  |  |  |  |  |  |  |
| **HP Class I** | 0.75 ± 0.07 | 0.92 ± 0.04 | 0.76 ± 0.11 | 0.46 ± 0.09 | 0.75 ± 0.05 | 41 ± 5 | 4 ± 2 | 11 ± 3 | 14 ± 4 |
| **HP Class II** | 0.96 ± 0.03 | 0.86 ± 0.03 | 0.44 ± 0.13 | 0.77 ± 0.16 | 0.85 ± 0.04 | 52 ± 5 | 8 ± 2 | 7 ± 2 | 2 ± 1 |
| **Milan** ^1^ | 0.80 ± 0.04 | 0.82 ± 0.04 | 0.35 ± 0.09 | 0.33 ± 0.09 | 0.70 ± 0.04 | 44 ± 5 | 10 ± 2 | 5 ± 2 | 11 ± 3 |
| **UCSF** ^2^ | 0.91 ± 0.03 | 0.80 ± 0.04 | 0.16 ± 0.08 | 0.33 ± 0.16 | 0.75 ± 0.04 | 49 ± 5 | 13 ± 2 | 2 ± 1 | 5 ± 2 |
| **AFP SAMPLES** |  |  |  |  |  |  |  |  |  |
| **HP Class I** | 0.76 ± 0.08 | 0.95 ± 0.04 | 0.81 ± 0.14 | 0.40 ± 0.12 | 0.77 ± 0.07 | 27 ± 4 | 1 ± 1 | 5 ± 2 | 9 ± 3 |
| **HP Class II** | 0.97 ± 0.03 | 0.91 ± 0.04 | 0.47 ± 0.19 | 0.74 ± 0.23 | 0.89 ± 0.04 | 34 ± 4 | 4 ± 2 | 3 ± 1 | 1 ± 1 |
| **AFP Score** ^3^ | 0.86 ± 0.04 | 0.87 ± 0.04 | 0.31 ± 0.16 | 0.29 ± 0.14 | 0.77 ± 0.04 | 31 ± 4 | 5 ± 2 | 2 ± 1 | 5 ± 2 |
| **ArgScore** ^4^ | 0.83 ± 0.05 | 0.88 ± 0.04 | 0.40 ± 0.18 | 0.31 ± 0.14 | 0.76 ± 0.05 | 29 ± 4 | 4 ± 2 | 3 ± 1 | 6 ± 2 |
| **Warsaw** ^5^ | 0.95 ± 0.03 | 0.86 ± 0.04 | 0.19 ± 0.14 | 0.42 ± 0.30 | 0.83 ± 0.05 | 34 ± 4 | 6 ± 2 | 1 ± 1 | 2 ± 1 |
| **MT2.0** ^6^ | 0.93 ± 0.04 | 0.86 ± 0.04 | 0.23 ± 0.15 | 0.37 ± 0.24 | 0.82 ± 0.05 | 33 ± 4 | 5 ± 2 | 2 ± 1 | 3 ± 1 |
| **wALL** ^7^ | 0.84 ± 0.05 | 0.87 ± 0.04 | 0.31 ± 0.16 | 0.27 ± 0.13 | 0.76 ± 0.05 | 30 ± 4 | 5 ± 2 | 2 ± 1 | 6 ± 2 |

HP – retrained HepatoPredict, Milan – Milan criteria, UCSF – University of California San Francisco criteria, AFP – alpha-fetoprotein, ArgScore – Argentinian Score, Warsaw – Warsaw criteria, MT2.0 – metroticket 2.0 criteria, wALL – within all criteria, PPV – positive predictive value, NPV – negative predictive value, TP – true positives, FP – false positives, TN – true negatives, FN – false negatives.

**Figures**

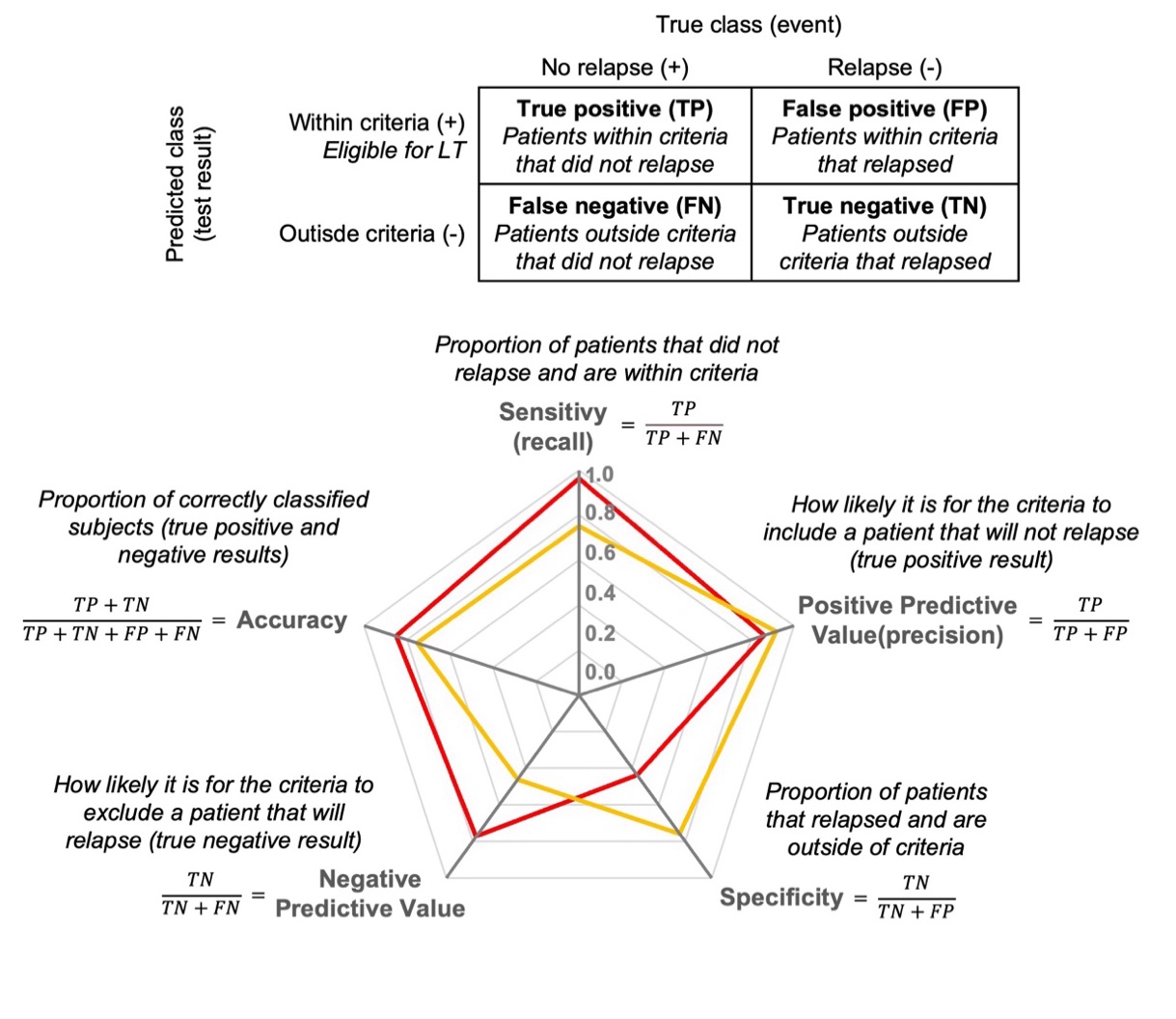

**Supplementary Figure 1 - Concepts and performance metrics employed in the study, including definitions and formulas.**

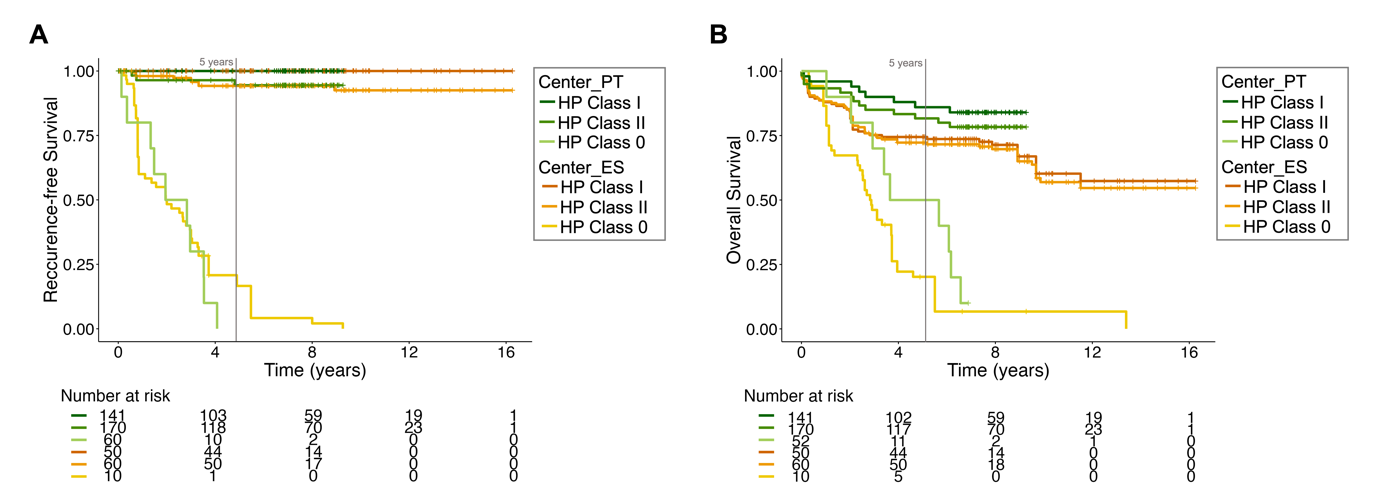

**Supplementary Figure 2 – Patients’ recurrence-free survival and overall survival in the different cohorts.** Comparison of the retrained HepatoPredict algorithm (HP) performance in patients from the Portuguese Center (Center_PT) – Hospital Curry Cabral, Lisbon, Portugal – and the Spanish Center (Center_ES) – Hospital Universitari i Politècnic La Fe, Valencia, Spain. Representation of the recurrence-free survival **(A)** and overall survival **(B)** curves and respective number of patients at risk at each time-point. HP Class I is a subset of HP Class II.

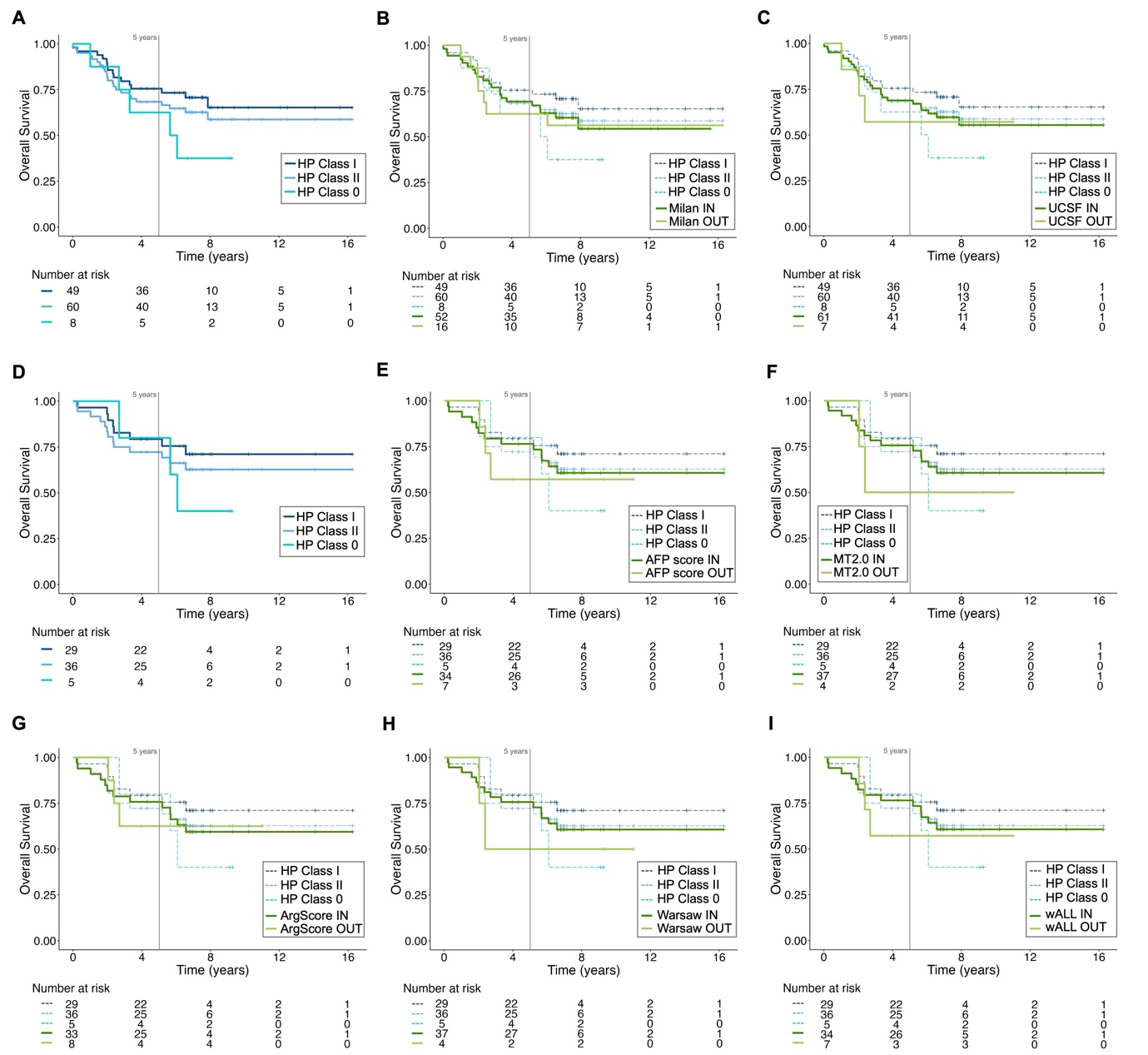

**Supplementary Figure 3 – Patients’ overall survival according to different criteria.** Overall survival curves (OS) of the most representative testing subset. The retrained HepatoPredict (HP) algorithm, Class I, II and 0 **(A)** was compared with Milan **(B)** and the University of San Francisco California (UCSF) **(C)** criteria, n = 68. OS was also calculated for the AFP samples subset of patients within the different HP classes **(D)** and compared with AFP-based criteria such as AFP score **(E)**, Metroticket 2.0 (MT2.0) **(F)**, Argentinian score (ArgScore) **(G)**, Warsaw **(H)**, and within all (wALL) criteria **(I)**, n = 41. For each criteria patients that are eligible (IN) and noneligible (OUT) for LT are represented. HP Class I is a subset of HP Class II.

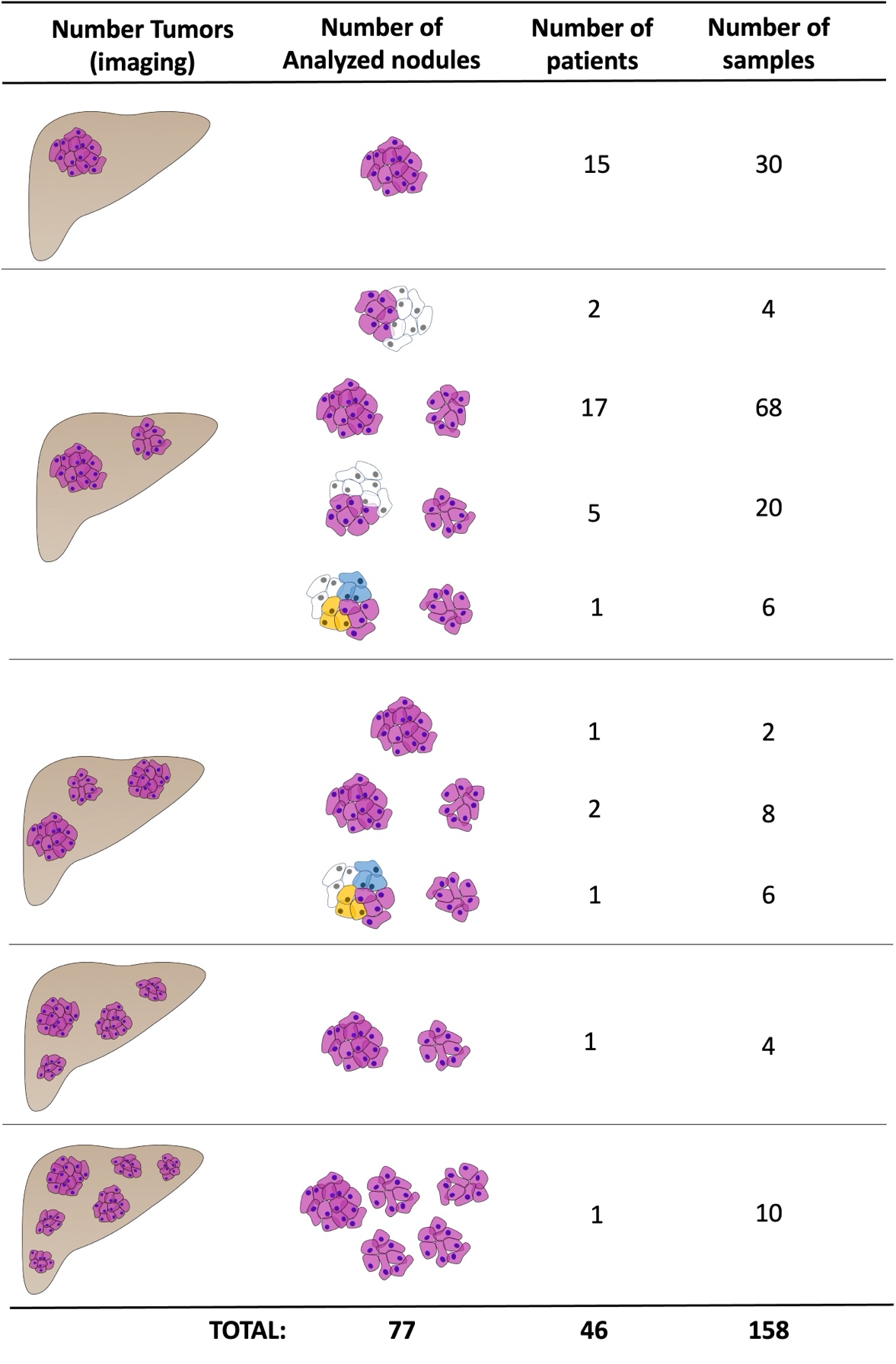

**Supplementary Figure 4 – Representation of dataset 2.** Dataset 2 was composed of 46 patients diagnosed with HCC and submitted to LT, from which it was possible to isolate 158 independent tumor areas from 77 nodules. The first column – *Number of Tumors (imaging)* – represents the number of tumors identified pre-LT and used as a variable for the HP algorithm. The second column – *Analyzed nodules –* represents the nodules with available tissue. Different colors within each nodule illustrate tissue heterogeneity. The total number of nodules, patients and samples are depicted.

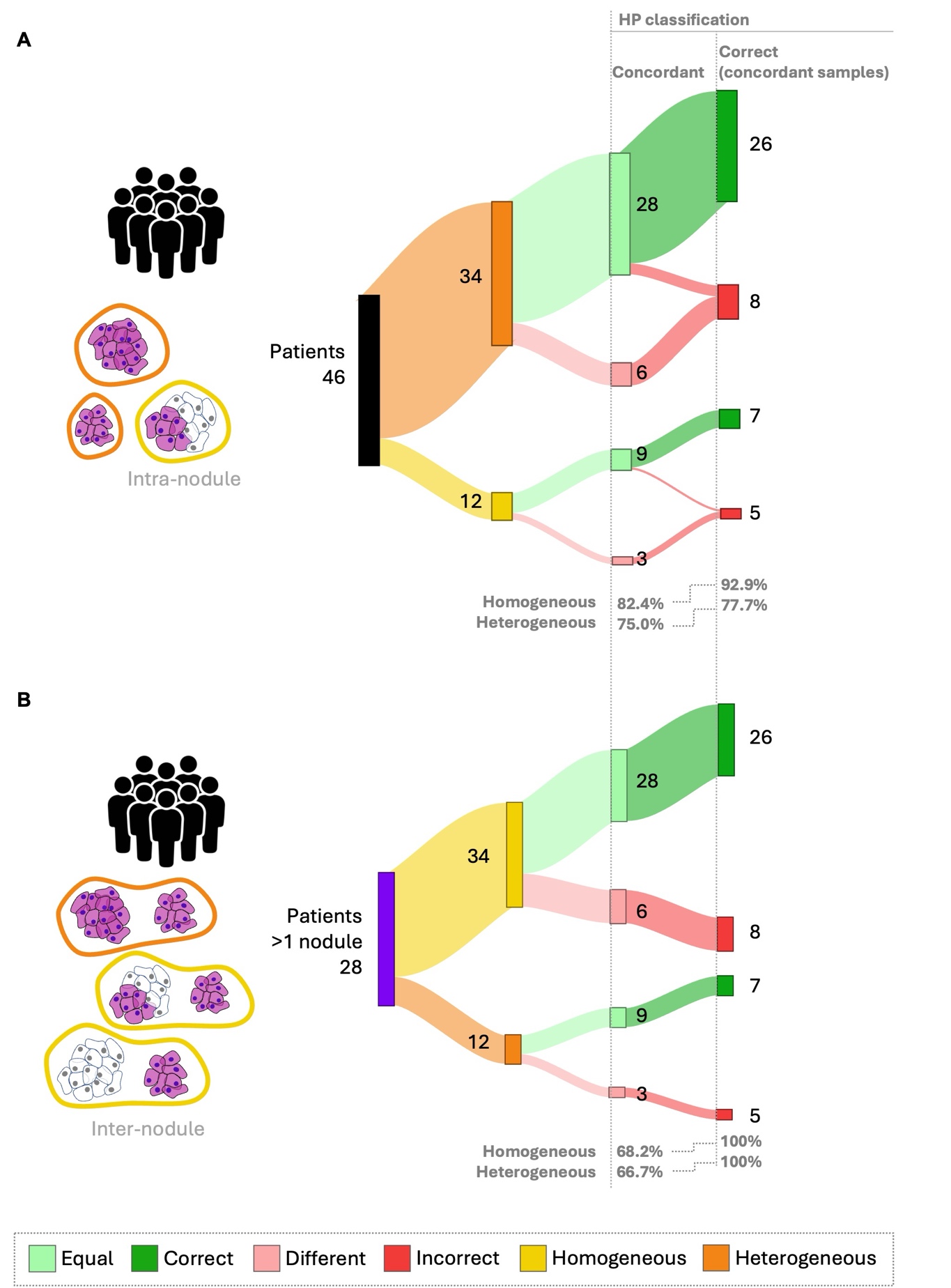

**Supplementary Figure 5 – HepatoPredict performance in the context of intra-nodule and inter-nodule heterogeneity – concordance and correct prediction in concordant samples.** To evaluate the impact of intra-nodule and inter-nodule heterogeneity on HP performance, at least two samples of each nodule were collected and characterized regarding the concordance of their HP assay results specifically, whether these samples from the same nodule received the same HP classification. The performance of HP, defined as its ability to produce correct prognoses, was evaluated using only the HP concordant samples. The analysis was performed for homogeneous and heterogeneous patients, based on intra-nodule (**A**) and inter-nodule (**B**) heterogeneity. The number (N) and percentage (%) are indicated for nodule/patients showing concordant vs. different HP results and for concordant samples. Any instance where the HP algorithm produced discordant results was automatically classified as incorrect.

**References:**

1.	Mazzaferro V, Regalia E, Doci R, Andreola S, Pulvirenti A, Bozzetti F, et al. Liver transplantation for the treatment of small hepatocellular carcinomas in patients with cirrhosis. N Engl J Med. 1996 Mar 14;334(11):693–702.

2.	Yao FY, Xiao L, Bass NM, Kerlan R, Ascher NL, Roberts JP. Liver transplantation for hepatocellular carcinoma: Validation of the UCSF-expanded criteria based on preoperative imaging. American Journal of Transplantation. 2007 Nov;7(11):2587–96.

3.	Duvoux C, Roudot-Thoraval F, Decaens T, Pessione F, Badran H, Piardi T, et al. Liver transplantation for hepatocellular carcinoma: A model including α-fetoprotein improves the performance of milan criteria. Gastroenterology. 2012;143(4).

4.	Piñero F, Marciano S, Anders M, Ganem FO, Zerega A, Cagliani J, et al. Identifying patients at higher risk of hepatocellular carcinoma recurrence after liver transplantation in a multicenter cohort study from Argentina. Eur J Gastroenterol Hepatol. 2016 Mar 4;28(4):421–7.

5.	Grąt M, Wronka KM, Stypułkowski J, Bik E, Krasnodębski M, Masior Ł, et al. The Warsaw Proposal for the Use of Extended Selection Criteria in Liver Transplantation for Hepatocellular Cancer. Ann Surg Oncol. 2017 Feb 1;24(2):526–34.

6.	Mazzaferro V, Sposito C, Zhou J, Pinna AD, De Carlis L, Fan J, et al. Metroticket 2.0 Model for Analysis of Competing Risks of Death After Liver Transplantation for Hepatocellular Carcinoma. Gastroenterology. 2018 Jan 1;154(1):128–39.

7.	Piñero F, Costentin C, Degroote H, Notarpaolo A, Boin IF, Boudjema K, et al. AFP score and metroticket 2.0 perform similarly and could be used in a “within-ALL” clinical decision tool. JHEP Reports. 2023 Feb 1;5(2).

8.	Gonçalves-Reis M, Proença D, Frazão LP, Neto JL, Silva S, Pinto-Marques H, et al. Analytical validation and algorithm improvement of HepatoPredict kit to assess hepatocellular carcinoma prognosis before a liver transplantation. Pract Lab Med. 2024 Mar;39:e00365.
